# Interoperability of standardised electronic healthcare records facilitates transfer learning of clinical language models

**DOI:** 10.1101/2025.06.12.25329419

**Authors:** Elizabeth Remfry, Rafael Henkin

## Abstract

**Objectives:** To investigate whether standardising electronic healthcare record (EHR) data to the Observational Medical Outcomes Partnership (OMOP) common data model improves cross-dataset interoperability and enables transfer learning of clinical language models for prediction tasks.

**Materials and Methods:** We used two UK datasets: Clinical Practice Research Datalink (CPRD) Aurum, restricted to London, and CPRD GOLD, covering all of England. Each dataset was independently mapped to OMOP standard concepts. A BERT-based transformer was pretrained on GOLD and fine-tuned on both datasets separately to predict 30-day emergency hospital readmission. An ablation study assessed the contribution of the three largest OMOP domains.

**Results:** OMOP mapping was successful for 95.6% of CPRD GOLD and 86.2% of Aurum records. Models fine-tuned on Aurum achieved comparable or superior performance (AUC: 0.93, 95% CI 0.93–0.93) than models fine-tuned on GOLD (AUC: 0.91, 95% CI 0.91–0.92). Ablation results diverged between datasets: GOLD performed better without measurements, while Aurum performed better without drugs.

**Discussion:** OMOP standardisation enabled effective transfer learning across datasets even in the presence of meaningful demographic differences, including ethnic composition and socioeconomic status. Divergence in ablation findings suggests that structural and content differences between primary care datasets influence downstream model behaviour. Incomplete vocabulary coverage within clinical language models also introduced data loss, representing a practical limitation for real-world deployment.

**Conclusion:** OMOP standardisation supported interoperability and transfer learning for clinical prediction using language model with high predictive performance.

## BACKGROUND AND SIGNIFICANCE

Electronic healthcare records (EHR) use clinical codes from ontologies to describe diseases, medications, and other clinical occurrences. Within the same country, or even within the same care system, these coding ontologies differ considerably between primary and secondary care observations, drug prescriptions and surgical procedures [1]. This heterogeneity is an obstacles to analysis and to training and deploying statistical and machine learning models that perform across systems and countries.

Common data models (CDMs) address this by standardising both the structure and the vocabulary of clinical data. The Observational Medical Outcomes Partnership (OMOP) CDM is among the most widely adopted, providing a shared framework of data structures and vocabularies that facilitates the combination and comparison of datasets across institutions and countries [2,3]. Standardisation has supported multinational network studies [4,5], in which researchers apply the same analytical procedures to their local datasets and combine only the aggregate results. As the data can remain at each local site, this approach can help avoid many of the privacy and data protection barriers of sharing patient-level data. OMOP-mapped data has been used, for example, in areas such as pharmacoepidemiology, investigating trends in drug usage across countries [6] or the impact of drug shortages around the world [7]. Standardisation is particularly valuable where no single dataset holds enough data. In rare diseases, harmonising multiple sources can provide adequate sample sizes [8] and in emerging diseases, reusing code and phenotype definitions, allows research to begin quickly, as demonstrated during the COVID-19 pandemic in studies of COVID-19 and asthma across different continents [9].

The same properties that make analyses portable across sites, also allow prediction models to be shared between them, an example of transfer learning, whereby a model, or the knowledge it has learned, from one dataset is applied in another setting. A common data structure and a set of standardised codes give models a consistent input representation, so a model can be reused and externally validated at other sites without access to the original training data [10]. Reps et al. used the OMOP framework to evaluate a range of prognostic models that predict stroke in patients with atrial fibrillation across 9 datasets from different countries, rapidly providing evidence of external validation [11].

Clinical foundation models, extend this ideas from single purpose models to reusable general ones. These models are large-scale neural networks pretrained on patient-level data and applied to different tasks, such as predicting diagnoses, mortality and hospital readmission, rather than requiring a separate model for [12]. Guo et al. [13] demonstrated that foundation models pretrained on a dataset from one US hospital continued to perform well on prediction tasks even when transferred to external datasets and that models adapted to external datasets required considerably less local data to reach a performance that was comparable to models trained from scratch. To support transfer of models across locations, Wornow et al. [14] released a reusable clinical foundation model trained on OMOP-standardised structured EHR data, which can be applied to other OMOP formatted datasets and applied to a variety of downstream clinical tasks.

Together, these studies highlight that transfer of models is feasible when data is standardised, yet little attention has been given to the impact of the standardisation process on the transferability. Mapping to a common data model often merges or discards some information [1] and it remains unclear how well the mapped representation preserves the information needed for prediction, or which of its components carry the signal that transfers between datasets. This is particularly salient for clinical language models, which require the use of a tokenizer built from fixed vocabulary of codes defined during pretraining and typically consisting of codes present in the original dataset. Codes absent from the pretraining data cannot be interpreted in a new dataset, so the success of the transfer may depend on how well the codes in the new dataset are covered by those the model has seen. Standardisation may reduce this problem by mapping different source codes onto shared standard concepts, although how far it does so has not been established.

In this study, we examine standardisation and transfer learning together, using emergency hospital readmission as a test case. Emergency hospital readmission, typically defined as unplanned readmission within 30 days of discharge, can be derived across different datasets that provide admission and discharge data and is a common benchmark for clinical language models [12]. We use two datasets drawn from the same national health system but differing markedly in population and underlying coding ontologies. We first assess the fidelity of the OMOP mapping for CPRD GOLD and CPRD Aurum, we then evaluate transfer learning by applying a clinical foundation model pretrained on CPRD GOLD to an external dataset, CPRD Aurum, and finally we use ablations by clinical domain to identify which components of the standardised representation carry the transferred signal.

## OBJECTIVES

The overall objective of this work is to investigate how standardising EHR data to the OMOP vocabularies supports the transfer of a clinical foundation model between two datasets. Specifically, we aim to assess the fidelity of the OMOP mapping to both CPRD Aurum and GOLD and to evaluate the transfer of foundation models pretrained on CPRD GOLD to CPRD Aurum for predicting emergency 30-day hospital readmission.

## MATERIALS AND METHODS

### Study Population

We used two primary care datasets collected from the National Health Service (NHS) in England, CPRD GOLD and Aurum. CPRD is a real-world research database which collects pseudonymised electronic healthcare records from a network of over 2,400 general practitioners (GP). Datasets were linked to the Office for National Statistics (ONS) mortality data and Index of Multiple Deprivation (IMD) for area-level deprivation data.

The study population included individuals aged at least 16 years, registered with a general practice between 1/1/2010 and 31/12/2020 whose data met CPRD’s acceptable data standards and were eligible for linkage to Hospital Episode Statistics (HES) data. From the CPRD Aurum dataset, we selected individuals registered to a GP based in London. From both CPRD Aurum and GOLD, we then selected a study population of individuals who had been diagnosed with at least two or more long-term conditions (LTCs) out of a code list of 204 [15] within our study period. From this cohort, we then extracted individuals who had experienced at least 1 hospitalisation between 1/1/2005 and 31/12/2019. This cutoff was chosen because it marks a significant shift in data collection patterns in the NHS [16].

### Outcome definition

The outcome was emergency readmission within 30 days of discharge. For each patient, the first admission after 1/12/2005 was the index admission. Cases were patients with a subsequent unplanned admission occurring at least 1 day after discharge and within 30 days of discharge. Controls were patients with no admission within 30 days, or whose admission was not an emergency, this included both elective (planned) and maternity admissions. Patients were excluded if they died during the index admission or were censored within 30 days of discharge, due to death, de-registration or practice’s last data contribution date.

### OMOP mapping procedure

#### Coding ontologies in CPRD

CPRD GOLD uses Read v2 codes for primary care events and Gemscript, a proprietary coding dictionary that includes the NHS Dictionary of Medicines and Devices (dm+d) for prescriptions. CPRD Aurum map to SNOMED CT UK Edition for primary care and dm+d for prescriptions. Within HES data, hospital diagnoses and procedures are coded using ICD-10 and Classification of Interventions and Procedures (OPCS-4), respectively.

#### Vocabularies and code standardisation

The OMOP CDM supports multiple clinical coding systems from health systems worldwide, collectively referred to as vocabularies within the OMOP ecosystem. For example, SNOMED CT is one of the medical vocabularies supported by OMOP. A vocabulary can contain both standard and non-standard concepts. A standard concept is considered the core representation of a medical event, so several non-standard concepts describing the same event can map to one standard concept. OMOP vocabularies and their mappings can be downloaded from the Observational Health Data Sciences and Informatics (OHDSI) Vocabularies Repository [17]. We downloaded the default set of vocabularies preselected in the OHDSI Athena browsing tool, supplemented with UK-specific vocabularies not preselected by default.

#### Mapping procedure

We defined Read, SNOMED CT, dm+d, ICD-10 and OPCS-4 as *source* vocabularies and used the concept relationship table to map (1) source codes to OMOP concept IDs, (2) non-standard concept IDs in *source* vocabularies to standard concept IDs in *target* vocabularies, and (3) standard concept IDs to the clinical codes they represent. For example, the Read v2 code “246n.00” for “baseline blood pressure” has an OMOP concept ID 45773921. This *source* concept ID in turn has an equivalence relationship with *target* concept ID 45770407, which is the SNOMED standard concept code 928021000000108 for the same event. We assessed the proportion of rows and unique codes that could not be mapped at each step, the reduction in unique concepts by dataset and table, and the distribution of domains and vocabularies after mapping.

### Model

#### Model input

Each patient’s records were represented as a chronologically ordered sequence of OMOP standard concept IDs, using all primary and secondary care data (Figure 1). We used OMOP concept IDs as they are unique between vocabularies. Patients with sequences of three or fewer concepts were excluded.

**Figure 1.**
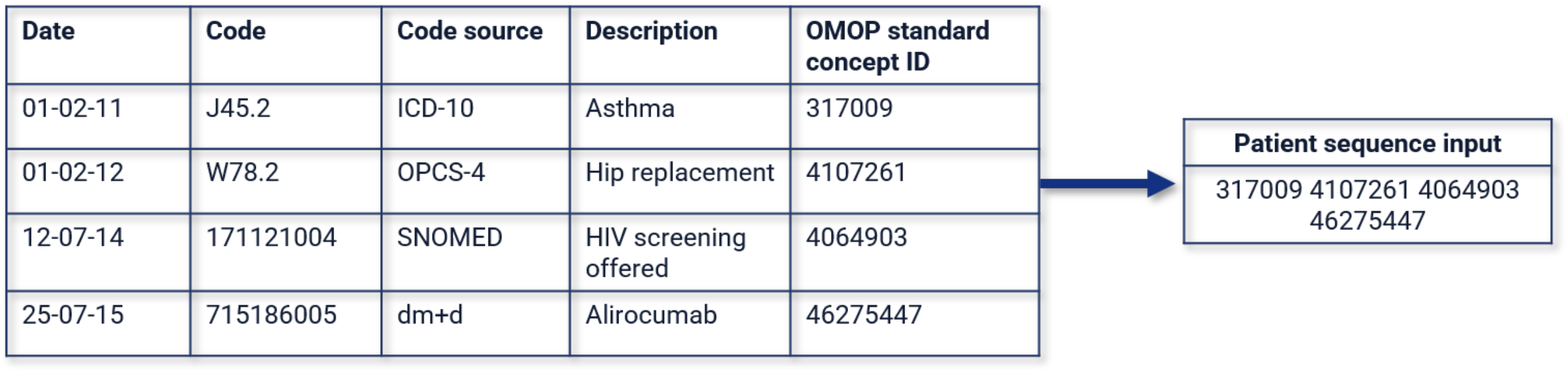
Mapping and format approach for input data for clinical language models

For pretraining, we utilised all data within a patient sequence, from date of entry into the cohort (1/01/2005) until death, deregistration or end of study (31/12/2020). For fine-tuning, only data up until the index hospital admission date was used.

CPRD GOLD was split by patient into 70% for pretraining and 30% for fine-tuning. The pretraining portion was further split into 95% for training and 5% for validation. The fine-tuning data from both CPRD GOLD and Aurum were split 70% training, 15% validation and 15% testing.

Hyperparameters were defined via Bayesian optimisation with Optuna [18]. For finetuning models Area Under the Receiver Operating Characteristic (AUROC) was used for optimisation. Hyperparameter search was conducted on a stratified 15% subset of each finetuning training set, which was withheld from the final model training.

#### Tokenizer

We trained a WordLevel tokenizer from scratch on the CPRD GOLD pretraining dataset so that each concept ID was represented by a single token, with a maximum token vocabulary of 20,000 and minimum frequency of 2. The same tokenizer was used for all models.

#### Pretraining

We pretrained an encoder only BERT-based transformer model from scratch on 1,051,302 individuals from CPRD GOLD (PT-GOLD) using masked language modelling, with 15% of tokens randomly masked. The model had 12 layers, a hidden size of 768, 12 attention heads and an intermediate size of 3072, totalling approximately 101 million parameters. Sequences were padded or truncated from the left to a maximum length of 512 tokens, retaining each individual’s most recent events.

#### Finetuning, baseline and transferability

For fine-tuning, a classification head was added to PT-GOLD, and all parameters were updated. We finetuned two models, one on CPRD GOLD (FT-GOLD) and the other on CPRD Aurum (FT-Aurum). Given the low frequency of cases (individuals with emergency readmission) we applied inverse frequency class weighting to the binary cross-entropy loss, upweighting positive cases by the ratio of negative to positive examples.

To isolate the effect of pretraining, we fine-tuned a control model on Aurum from a randomly initialised encoder (RI-Aurum), identical to FT-Aurum in architecture, tokenizer, training data and class weighting. To assess transferability, we applied FT-GOLD directly to the Aurum test set with no further training (INF-Aurum), reusing its weights and tokenizer to mimic external validation.

#### Evaluation

We assessed variation in model performance using a bootstrap resampling method with 1000 iterations to calculate 95% confidence intervals (CIs). We report the area under the receiver operating characteristic curve (AUROC) and the area under the precision-recall curve (AUPRC), with 95% confidence intervals from bootstrap resampling over 1,000 iterations. F1, precision and recall are reported in the Supplementary Materials. Experiments ran on one A100 GPU on Queen Mary’s Apocrita HPC facility supported by QMUL Research IT [19].

#### Ablation of input data

OMOP CDM structures concepts across over 30 domains including drugs, measurements, observation and conditions. However, the allocation of a concept can differ depending on the source dataset, for example a blood pressure reading in Aurum is classified as a *Measurement* as it contains a numerical value, whilst in GOLD a blood pressure reading is mapped to *Conditions and Observations*.

To assess the contribution of each OMOP domain, we remove one domain at a time from the input sequences at the fine-tuning stage for both FT-GOLD and FT-Aurum and assess performance on AUROC and AUPRC. Given that 90% of the data sits across drugs, measurements, observations and conditions we focus on these domains only. Given the high level of semantic overlap between Observations and Conditions we treat these as a single group.

## RESULTS

### Study population

Table 1 shows the demographic characteristics of patients across the three dataset splits (GOLD pretraining, GOLD and Aurum readmission). Emergency readmission occurred in 15.4%, 15.5% and 16.4% of patients across cohorts, respectively. The Aurum cohort was younger, with a median age of 48 years compared to 54 years in GOLD and higher levels of deprivation, which is consistent with variations in geographical differences.

**Table 1:**
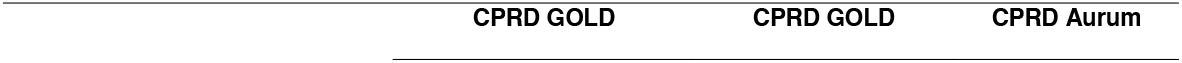
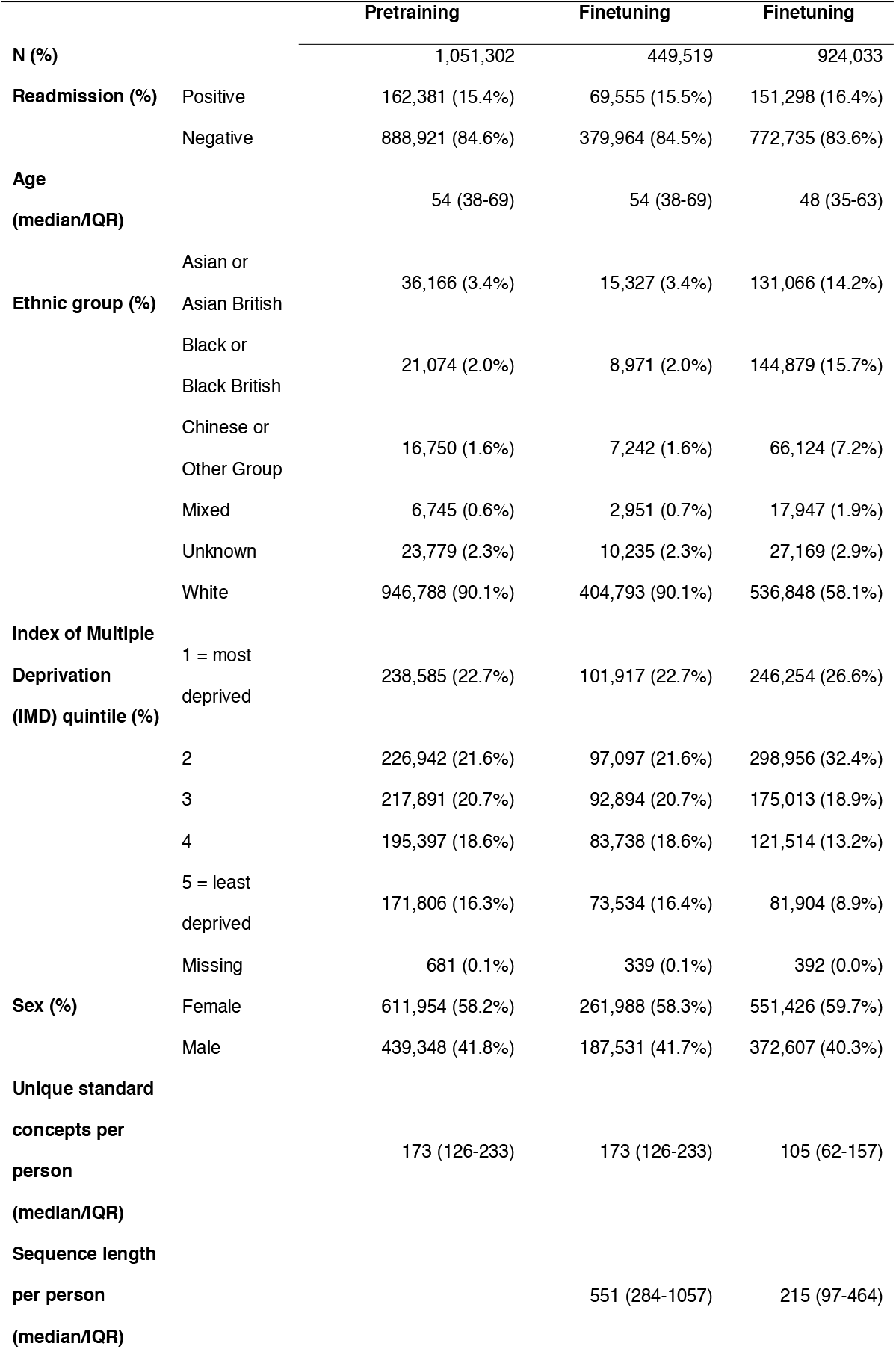
Demographic characteristics of patients in the three datasets.

### Concept mapping

We applied the OMOP mapping to the full CPRD GOLD and Aurum datasets before splitting into pretraining and fine-tuning cohorts. At least 94% of rows mapped to a standard concept in 7 of the 8 source data tables (see Figure 2). Only the primary care table in CPRD Aurum had a lower success rate where 80% of rows mapped, 8.7% mapped to a non-standard concept or missing vocabulary, and 11% of rows were not found in the OMOP version of SNOMED CT. The raw counts can be found in Supplementary Table S1.

**Figure 2.**
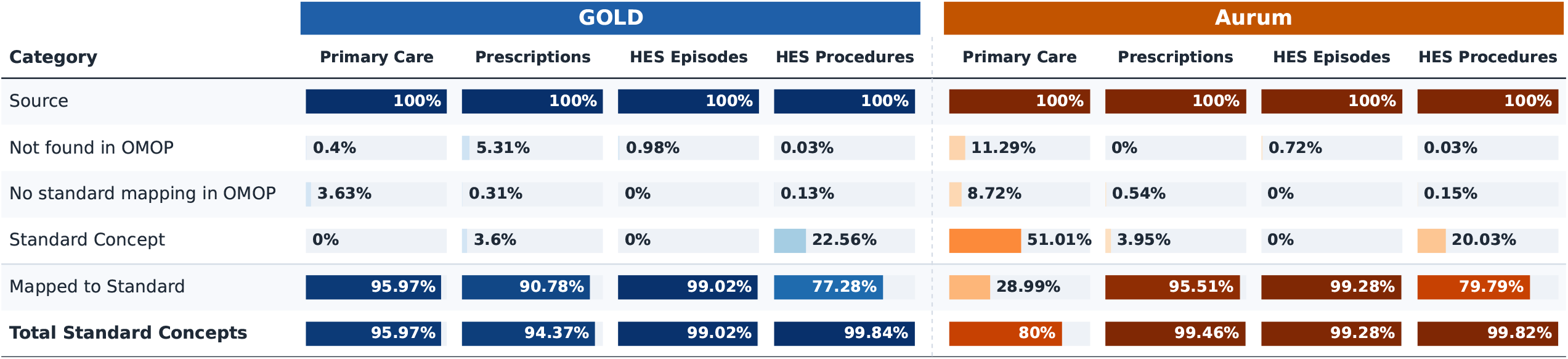
Percentage of concepts in each step of the mapping pipeline, per table in each CPRD Dataset, relative to the original tables.

Mapping also reduced the number of unique concepts. In primary care, unique concepts fell by 36% in GOLD (from 80,000 to 51,000) and by 28.6% in Aurum (from 73,000 to 52,000). Prescription data showed different reductions for the two datasets, 27.5% in GOLD and 3% in Aurum, although both are based on dm+d derived coding.

### Domain results

After mapping the Drug domain was the most common domain in GOLD, accounting for 34.3% of rows, followed by Measurement at 28.3%, whereas in Aurum, Measurement was the most common, accounting for 36.9%, followed by Drug, at 30.1%. In both datasets, the combination of Observation and Condition had 27.3% and 25.4% rows, respectively. SNOMED CT was the most common vocabulary in both datasets after mapping, with 62.1% and 66.6% in GOLD and Aurum, respectively.

### Tokenizer coverage

The tokenizer vocabulary, capped at 20,000 tokens, covered 99.53% of concept occurrences in the CPRD GOLD pretraining corpus, corresponding to 19,995 of 84,558 distinct concept IDs (23.65%). For the CPRD GOLD finetuning dataset, the tokenizer vocabulary covered 99.53% of concept occurrences, which covers 19,974 of 61,230 distinct concept IDs (32.62%). Whilst for CPRD Aurum, the vocabulary of the tokenizer covered 91.27% of the occurrences, corresponding to 16,966 of 57,928 distinct concept IDs (29.29%). This reflects the highly skewed frequency distribution of clinical codes, where the majority of codes are not used frequently.

### Model performance

Table 2 shows AUROC and AUPRC for all models, additional metrics F1, recall and precision are reported in Supplementary Table S2. Fine-tuned models performed well across both datasets. FT-GOLD achieved an AUROC of 0.913 (95% CI: 0.910 - 0.916) on the GOLD test set and FT-Aurum 0.927 (0.925 - 0.928) on the Aurum test set.

**Table 2.** Metrics for model performance.

|  | **AUROC** | **AUPRC** |
| --- | --- | --- |
| **FT-GOLD** | 0.913 (0.910-0.916) | 0.794 (0.787 - 0.800) |
| **FT-Aurum** | 0.927 (0.925-0.928) | 0.812 (0.808 - 0.817) |
| **RI-Aurum** | 0.885 (0.883 - 0.888) | 0.675 (0.668 - 0.681) |
| **INF-Aurum** | 0.911 (0.909 - 0.914) | 0.757 (0.752 - 0.762) |

FT-GOLD: Base model pretrained on GOLD and finetuned on GOLD; FT-Aurum: Base model pretrained on GOLD and finetuned on Aurum; RI-Aurum: Base model with randomly initialised weights and finetuned on Aurum; INF-Aurum: Base model pretrained and finetuned on GOLD, inference tested on Aurum.

The remaining comparisons were evaluated on the same CPRD Aurum test set. RI-Aurum, finetuned on a randomly initialised encoder achieved an AUROC of 0.885 (0.883–0.888). FT-Aurum, differing in that its encoder was pretrained on CPRD GOLD, exceeded this by +0.042 AUROC, indicating that pretraining contributed to downstream performance even though it was conducted on a different database.

FT-GOLD, a model pretrained and finetuned on GOLD only, transferred successfully at inference only when applied to Aurum (INF-Aurum), with an AUROC of 0.911 (0.909 - 0.914). Fine-tuning further on Aurum (FT-Aurum) gained a further +0.016 AUROC over inference alone.

### Ablation

Removing observations and conditions reduced performance in both datasets, from an AUROC of 0.913 (0.910 to 0.916) to 0.897 (0.894 to 0.901) in FT-GOLD and from 0.927 (0.925 to 0.928) to 0.888 (0.886 to 0.891) in FT-Aurum **(Table 3)**. Removing drugs or measurements, left performance unchanged or improved it. FT-GOLD performed best with measurements removed (AUROC 0.929, 0.926–0.932), while FT-Aurum performed best with drugs removed (AUROC 0.931, 0.929–0.933). Additional metrics F1, recall and precision are reported in Supplementary Table S3.

**Table 3.**
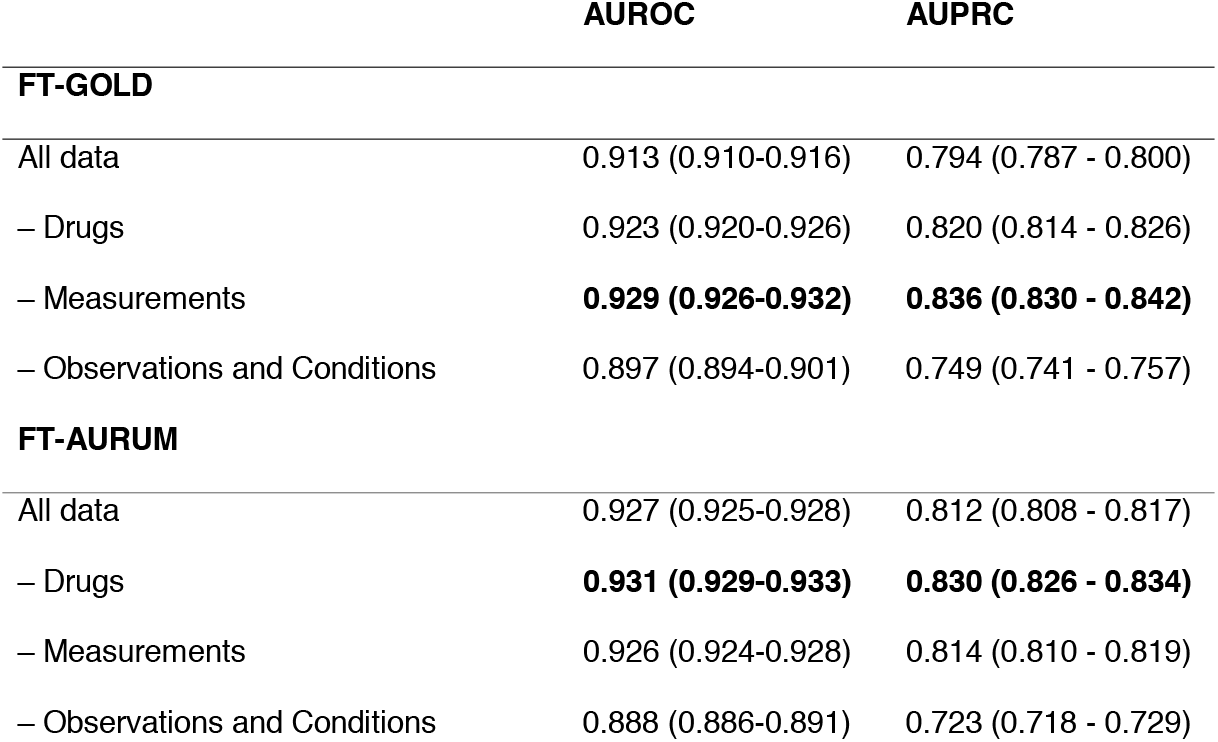
Model performance across ablation studies.

## DISCUSSION

In this study we mapped CPRD GOLD and Aurum to the OMOP standard vocabularies and used the shared concept space to transfer a clinical language model from one dataset to the other.

Mapping to OMOP concepts was largely successful in both datasets, which is in line with previous CPRD conversions [20–22]. However, for Aurum, although our results mirrored previous research which found that the largest table in OMOP was measurements, overall only 51.01% of source concepts were already standard in this work, as compared to 60.1% previously reported [20]. This may be due to differences in our study population, as our study was not conducted on the entire CPRD Aurum population.

We found differences between the datasets in the overall level of codes mapped. The two primary care datasets had two distinct source vocabularies, CPRD Aurum utilised SNOMED CT, which is standard in OMOP, and achieved 80% of codes mapped. Whilst CPRD GOLD used Read codes, which were not standard, but mapped with very high levels of success (96%). We suggest that this divergence (80% versus 96%) may be due to differences in the UK edition of SNOMED CT as compared to other international editions of SNOMED CT. UK specific terms are not recognised as standard concepts, and therefore fail to map. Hospital data derived from the same coding ontologies (ICD-10, OPCS-4) in both datasets, therefore mapping results were similar. In GOLD, 99.84% of OPCS-4 mapped successfully, whilst in Aurum this was 99.83%.

A key element to this work is that we relied on standard OMOP concepts to harmonise the data. Other OMOP analyses have focused on standardisation based on the common data model and internal table structures, whereas our interest was in ensuring that datasets were interoperable simply at the coding level to facilitate transfer learning and sharing of foundation models. Mapping to OMOP using standard concepts only meant that the data in this study was compressed. Multiple source codes frequently mapped onto a single OMOP concept, reducing unique primary care concepts by 36% in GOLD and 28.6% in Aurum, and high-volume administrative codes, such as “Attachment” (5% of Aurum rows) and “Awaiting clinical code migration” (4% of all rows), were removed entirely because they had no standard concept.

Using sequences of standard OMOP concepts to represent patient trajectories, a model pretrained on GOLD transferred to Aurum with no update to its weights, achieving an AUROC of 0.911 (95% CI 0.909–0.914), and improved further after fine-tuning on Aurum (AUROC 0.927, 95% CI 0.925–0.928). Both exceeded a model trained on Aurum from a randomly initialised encoder (AUROC 0.885, 95% CI 0.883–0.888), indicating that pretraining on a differently coded dataset still contributed to downstream performance. These results held despite substantial demographic differences between the datasets in age, ethnicity and deprivation, suggesting some potential for models to be reused across settings even where standardisation has not been designed with transfer specifically in mind.

These findings are consistent with other work on transfer across clinical foundation models. Guo et al. found that a model pretrained at one US hospital and fine-tuned on both its own data and an external children’s hospital dataset outperformed models trained from scratch on a 30-day readmission task [13]. Hur et al. similarly reported that pretraining across datasets improved downstream performance, particularly when only a small amount of target-dataset data was available for fine-tuning [23,24].

The ablation results show that the OMOP domains retained in the input affect performance, and that their contribution differs by dataset. Removing observations and conditions reduced performance in both fine-tuned models, suggesting these domains carry information relevant to readmission risk. In contrast, removing drugs improved performance in FT-Aurum and removing measurements improved performance in FT-GOLD. These findings may also reflect how domains are assigned. We found that clinically comparable concepts, such as blood pressure readings, can be allocated to different OMOP domains in GOLD and Aurum, consistent with prior validation work noting those discrepancies arise from codes being mapped to different tables [22]. This means a given domain does not necessarily represent the same clinical content across the two datasets, which is a source of divergence separate from coding system differences alone.

There are various limitations with this approach. The tokenizer was constructed from CPRD GOLD alone, and covered 99.5% of concepts in GOLD but 91.2% in Aurum. Future work should explore training tokenizers across datasets or use tokenisation strategies that represent unseen codes rather than discarding them, such as MedTok [25] or MedRep, which derives concept embeddings from ontology structure and textual definitions [26]. Our pragmatic vocabulary download strategy means that some codes in both datasets probably went unmapped because the relevant vocabulary was not retrieved. We also did not quantify data loss at the patient level; future work should establish where, and for whom, information is lost in conversion to OMOP as this may have equity implications given the demographic differences between the two databases.

Finally, standardisation is a step towards model interoperability, but does not guarantee it, as OMOP standard concepts are drawn from coding systems from multiple countries. For example, the EHRSHOT model by Wornow et al. [14] was trained on OMOP-compatible data but predominantly uses LOINC codes for laboratory measurements. These measurements are typically recorded as standard concepts in SNOMED CT in UK datasets, which means that there is little data and tokenizer vocabulary overlap within the EHRSHOT model for UK datasets. Mapping from LOINC to SNOMED and vice-versa is a task separate from OMOP standardisation and requires considerable domain knowledge.

## CONCLUSION

Standardisation of EHR data to OMOP allowed a model pretrained on one UK primary care dataset, CPRD GOLD, to be reused on another built on a different coding system, CPRD Aurum, with no further training (AUROC 0.911), performing close to a model fine-tuned on Aurum (0.927) and ahead of a randomly initialised model (0.885). This held despite substantial demographic differences between the two populations and differences in OMOP mapping success, domain composition and tokenizer coverage all differing between datasets.

Mapping to standard OMOP concepts was largely successful across both datasets, though this compressed the data, merging multiple source codes into single concepts and removing high-volume administrative codes with no standard equivalent. These results indicate that OMOP standardisation, applied pragmatically at the concept level rather than through the full CDM structure, can support the reuse of clinical prediction models across settings, without training separate models from scratch, though the extent to which these findings generalise beyond two datasets from the same national health system remains to be tested.

## Data Availability

The data underlying this article is provided by the UK CPRD electronic health record database, which is only accessible to researchers with protocols approved by CPRD Research Data Governance.

## ACKNOWLEDGMENTS

This study is based in part on data from the Clinical Practice Research Datalink obtained under licence (21_000345) from the UK Medicines and Healthcare products Regulatory Agency.

## DATA AVAILABILITY

The Clinical Practice Research Datalink (CPRD) does not allow the sharing of patient-level data. The data were used under license for the current study. The list of long-term conditions and respective code lists used in this study are available at: https://github.com/Fabiola-Eto/MULTIPLY-Initiative.

