## Supplement for "Interoperability of standardised electronic healthcare records facilitates transfer learning of clinical language models"

**Supplementary Table S1.** Mapping of source codes to standard vocabularies, by data type, in CPRD GOLD and CPRD Aurum.

|  | **CPRD GOLD** | | **CPRD Aurum** | |
| --- | --- | --- | --- | --- |
|  | **n** | **%** | **n** | **%** |
| **Primary care (Read codes / SNOMED CT)** | | | | |
| Rows with valid codesᵃ | 913,403,805 |  | 853,175,026 |  |
| Unmapped | 3,624,974 | 0.4 | 96,283,560 | 11.3 |
| Non-standard / missing vocabulary | 33,179,841 | 3.6 | 74,370,571 | 8.7 |
| Directly mapped to standard | 0 | 0.0 | 435,214,888 | 51.0 |
| Mapped to standard via relationship | 876,598,990 | 96.0 | 247,306,007 | 29.0 |
| **Total rows mapped to standard** | **876,598,990** | **96.0** | **682,520,895** | **80.0** |
| **Prescriptions (dm+d)** | | | | |
| Rows with valid codesᵃ | 573,808,585 |  | 333,869,579 |  |
| Unmapped | 30,477,715 | 5.3 | 0 | 0.0 |
| Non-standard / missing vocabulary | 1,800,473 | 0.3 | 1,792,004 | 0.5 |
| Directly mapped to standard | 20,632,505 | 3.6 | 13,193,858 | 3.9 |
| Mapped to standard via relationship | 520,897,891 | 90.8 | 318,883,717 | 93.4 |
| **Total rows mapped to standard** | **541,530,396** | **94.4** | **332,077,575** | **97.3** |
| **HES diagnoses (ICD-10)** | | | | |
| Rows with valid codesᵃ | 75,582,095 |  | 46,123,776 |  |
| Unmapped | 739,015 | 1.0 | 330,872 | 0.7 |
| Non-standard / missing vocabulary | 8 | 0.0 | 3 | 0.0 |
| Directly mapped to standard | 0 | 0.0 | 0 | 0.0 |
| Mapped to standard via relationship | 74,843,072 | 99.0 | 45,792,901 | 99.3 |
| **Total rows mapped to standard** | **74,843,072** | **99.0** | **45,792,901** | **99.3** |
| **HES procedures (OPCS-4)** | | | | |
| Rows with valid codesᵃ | 26,820,756 |  | 16,089,586 |  |
| Unmapped | 7,006 | <0.1 | 5,055 | <0.1 |
| Non-standard / missing vocabulary | 34,968 | 0.1 | 23,617 | 0.1 |
| Directly mapped to standard | 6,050,857 | 22.6 | 3,222,835 | 20.0 |
| Mapped to standard via relationship | 20,727,925 | 77.3 | 12,838,079 | 79.8 |
| **Total rows mapped to standard** | **26,778,782** | **99.8** | **16,060,914** | **99.8** |

Values are numbers of rows (n) and percentages of rows with valid codes (%). <0.1 indicates a non-zero value below 0.05%.

^a^ Percentages are not shown for rows with valid codes, as this is the denominator.

**Supplementary Table S2.** Additional metrics, precision, recall and F1 for model performance

|  | **Precision** | **Recall** | **F1** |
| --- | --- | --- | --- |
| **FT-GOLD** | .485 (.478 - .492) | .813 (.806 - .820) | .608 (.601 - .614) |
| **FT-Aurum** | **.**520 (.514 - .524) | .841 (.836 - .845) | .642 (.638 - .646) |
| **RI-Aurum** | .438 (.433 - .442) | .862 (.857 - .866) | .581 (.576 - .585) |
| **INF-Aurum** | .407 (.403 - .412) | .837 (.832 - .842) | .548 (.543 - .552) |

**Supplementary Table S3.** Additional metrics, precision, recall and F1 for ablation

|  | **Precision** | **Recall** | **F1** |
| --- | --- | --- | --- |
| **FT-GOLD** | .485 (.478-.492) | **.813 (.806-.820)** | .608 (.601-.614) |
| **Without Drugs** | .622 (.614-.629) | .776 (.768-.784) | .690 (.684-.697) |
| **Without Measurements** | **.680 (.671-.689)** | .764 (.756-.772) | **.720 (.712-.727)** |
| **Without Observations and Conditions** | .647 (.638-.657) | .684 (.675-.693) | .665 (.658-.673) |
| **FT-Aurum** | .520 (.514-.524) | **.841 (.836-.845)** | .642 (.638-.646) |
| **Without Drugs** | **.629 (.623-.634)** | .798 (.793-.803) | **.703 (.699-.708)** |
| **Without Measurements** | .547 (.542-.552) | .828 (.823-.833) | .659 (.654-.663) |
| **Without Observations and Conditions** | .568 (.562-.574) | .715 (.710-.721) | .633 (.628-.638) |

Metrics in bold highlighted for best performance in each dataset
